# Neuronal alpha-Synuclein Disease Integrated Staging System performance in PPMI, PASADENA, and SPARK baseline cohorts

**DOI:** 10.1101/2024.02.14.24302818

**Authors:** Tien Dam, Gennaro Pagano, Michael C Brumm, Caroline Gochanour, Kathleen L Poston, Daniel Weintraub, Lana M. Chahine, Christopher Coffey, Caroline M. Tanner, Catherine M. Kopil, Yuge Xiao, Sohini Chowdhury, Luis Concha-Marambio, Peter DiBiaso, Tatiana Foroud, Mark Frasier, Danna Jennings, Karl Kieburtz, Kalpana Merchant, Brit Mollenhauer, Thomas J Montine, Kelly Nudelman, John Seibyl, Todd Sherer, Andrew Singleton, Diane Stephenson, Matthew Stern, Claudio Soto, Eduardo Tolosa, Andrew Siderowf, Billy Dunn, Tanya Simuni, Kenneth Marek, the Parkinson’s Progression Markers Initiative

## Abstract

The Neuronal alpha-Synuclein Disease (NSD) biological definition and Integrated Staging System (NSD-ISS) provide a research framework to identify individuals with Lewy body pathology and stage them based on underlying biology and increasing degree of functional impairment. Utilizing data from the PPMI, PASADENA and SPARK studies, we developed and applied biologic and clinical data-informed definitions for the NSD-ISS across the disease continuum. Individuals enrolled as Parkinson’s disease, Prodromal, or Healthy Controls were defined and staged based on biological, clinical, and functional anchors at baseline. Across the three studies 1,741 participants had SAA data and of these 1,030 (59%) were S+ consistent with NSD. Among sporadic PD, 683/736 (93%) were NSD, and the distribution for Stages 2B, 3, and 4 was 25%, 63%, and 9%, respectively. Median (95% CI) time to developing a clinically meaningful outcome was 8.3 (6.2, 10.1), 5.9 (4.1, 6.0), and 2.4 (1.0, 4.0) years for baseline stage 2B, 3, and 4, respectively.

We propose pilot biologic and clinical anchors for NSD-ISS. Our results highlight the baseline heterogeneity of individuals currently defined as early PD. Baseline stage predicts time to progression to clinically meaningful milestones. Further research on validation of the anchors in longitudinal cohorts is necessary.

## INTRODUCTION

We recently proposed a new research biological framework for Neuronal alpha-Synuclein Disease (NSD) and an integrated staging system (NSD-ISS)^1^ enabled by the development and validation of assays that can accurately detect misfolded neuronal alpha-synuclein (n-asyn) *in vivo*^2^. This biological definition is grounded on three key tenets: 1) a disease is defined biologically based on validated *in-vivo* biomarkers; 2) the disease can be diagnosed in absence of clinical manifestations; and 3) the same biology may result in different phenotypic presentations; thus, symptoms are a result of the disease process but do not define it. As such, diagnosis is based on disease-specific biomarkers, and symptoms are not necessary for diagnosis. This biological definition is a departure from traditional clinical diagnostic criteria of Parkinson’s disease (PD) and dementia with Lewy bodies (DLB)^3^ ^4^, which are biologically linked by the same aggregates of n-asyn found predominantly in neuronal cell bodies and neurites, but diverge based on the predominance of motor versus cognitive symptoms at initial clinical manifestation.

NSD is a unifying term that encompasses PD, DLB, and any other n-asyn driven clinical syndrome. We have further proposed the NSD-ISS which integrates the biological substrates of the disease, n-asyn (S) and dopaminergic dysfunction (D), with cognitive, other non-motor, or motor manifestations and functional impairment to define stages along the NSD continuum. The intent of the NSD-ISS is to provide an integrated biological and clinical framework to expand understanding of disease and advance biologically targeted therapeutic development.

The NSD-ISS proposed seven distinct stages: Stage 0 (presence of fully penetrant pathogenic variants in *SNCA* gene); Stage 1 (presence of n-asyn alone (Stage 1A) or in combination with dopaminergic dysfunction (Stage 1B), asymptomatic); Stage 2 (presence of n-asyn alone (Stage 2A) or in combination with dopaminergic dysfunction (Stage 2B), and subtle clinical signs/symptoms without functional impairment); and Stages 3-6 (presence of both n-asyn and dopaminergic dysfunction, and clinical signs/symptoms with progressively increasing severity of functional impairment).

Simuni et al. outlined the staging framework in a Position paper^1^. The objectives of this study were to 1) develop biologic and clinical criteria and thresholds, utilizing currently available clinical scales, to operationalize the NSD and NSD-ISS framework ; 2) apply these definitions across the disease continuum utilizing available data in three well characterized studies; and 3) evaluate time to onset of key clinical outcomes based on baseline NSD stage.

## RESULTS

Participant-level data from three independent studies were evaluated: Parkinson’s Progression Markers Initiative (PPMI), PASADENA and SPARK. Respective study aims and methodology have been published elsewhere^5–7^. All studies and recruitment materials were approved by institutional review boards or ethics committee at each site. Written informed consent was obtained from all participants before undergoing any study evaluations. Clinical trials were performed in accordance with the principles outlined in the Declaration of Helsinki and with Good Clinical Practice guidelines.

Briefly, the PPMI (NCT01141023) study is a multinational, prospective longitudinal observational study launched in 2010^5^ with three cohorts: clinically diagnosed early PD, prodromal, or non-manifesting carriers of genetic variants associated with PD, and healthy controls (HC). Individuals with PD were enrolled if they were within 2 years of diagnosis, Hoehn and Yahr (H&Y) stage 1–2, not on PD medications at the time of enrollment, and had an abnormal dopamine transporter (DAT) imaging scan with single-photon-emission computed tomography (SPECT). Inclusion criteria for the genetic PD cohort were the same, except for PD medications and diagnosis within 7 years were allowed. Prodromal participants had prodromal features associated with risk of PD, including severe hyposmia as measured by the University of Pennsylvania Smell Identification Test (UPSIT) based on internal population norms^8^ or REM sleep behavior disorder (RBD) confirmed by polysomnogram. Healthy controls were similar age- and sex individuals without known neurological signs or symptoms and normal DAT imaging. All PPMI participants undergo extensive clinical phenotypic and biological characterization annually that includes collection of cerebrospinal fluid (CSF) samples and DAT imaging. Details regarding the protocol and imaging data are posted online^9^. All participants undergo whole genome sequencing after recruitment, and participants with relevant genetic variants are analyzed accordingly.

PASADENA (NCT03100149) was a phase 2, multinational, double-blind, randomized controlled trial examining the efficacy and safety of prasinezumab in 316 individuals with early PD who received intravenous prasinezumab (1500 mg or 4500 mg) or placebo every 4 weeks for 52 weeks^6^.

SPARK (NCT03318523) was a phase 2, multinational, double-blind, randomized controlled trial that examined the efficacy and safety of cinpanemab in 357 individuals with early-stage PD were assigned to receive one of three doses (250 mg, 1250 mg, or 3500 mg) intravenous cinpanemab or placebo every 4 weeks for 52 weeks, after which placebo recipients switched to cinpanemab^7^.

Participants from all three studies underwent a series of clinical assessments described previously^5–7^. Relevant assessments included the Montreal Cognitive assessment (MoCA)^10^ and the Movement Disorders Society Unified Parkinson’s Disease Rating Scale (MDS-UPDRS) parts I (non-motor aspects or experiences of daily living), II (motor aspects or experiences of daily living), and III (motor examination; recorded in the off**-**state for treated participants)^11^. Lumbar punctures for CSF was required at baseline for PPMI and was collected in a subset of participants in both PASADENA and SPARK,.

The NSD-ISS Framework categorized individuals based on biologic (S, D, and G), clinical, and functional impairment anchors (Supplementary Table 1)^1^. See Supplementary Table 2 for the glossary of terms.

### S anchor

The presence of n-asyn was evaluated using a CSF α-synuclein (n-asyn) seed amplification assay (n-asyn SAA)^2,12–14^. Samples from the three studies followed the same sample processing procedures and were analyzed at a central laboratory using standardized assay conditions (Amprion). Each individual sample was analyzed in triplicate and determined to be either αSyn-SAA positive (S+) or negative (S-) according to a previously reported algorithm ^15^. Some PPMI participants only had CSF n-asyn SAA evaluated at a follow-up visit; in such cases, participants were considered S+ if they tested positive within 12 months (PD cohort) or 6 months (other cohorts) of baseline, and were considered S- if they tested negative at any follow-up visit.

### D anchor

The presence of dopamine dysfunction was evaluated in all 3 studies with DAT imaging using ^123^I-ioflupane. Images were processed at a central laboratory with a standardized reconstruction algorithm and image analysis workflow (PMOD for PPMI and PASADENA, Hermes Medical Solutions for SPARK) and were analyzed visually by experienced nuclear medicine experts unaware of the trial-group assignments. Visual interpretation of the scan was used as a criterion for enrollment into PPMI PD cohorts, PASADENA, and SPARK. Quantitative assessment of specific binding ratio (SBR) in striatal regions was calculated using previously developed methods^16^. Lowest SBR adjusted for age and sex was used to determine DAT deficit for NSD-ISS staging. Individual with a putamen SBR ≤75% were designated as D+. Based on this quantitative criterion, an individual with dopamine dysfunction by visual inspection could be D-. Three PPMI PD participants underwent VMAT-2 imaging with 18F AV133 (not DAT imaging) and were assumed to be D+ based on visual inspection only.

### G anchor

The criterion for Stage 0, defined strictly by G+ status was restricted to only fully penetrant pathogenic *SNCA* variants. Non-manifesting carriers of other relevant genetic variants who were S- were included in the at-risk category but were not considered NSD.

Process for defining anchors for clinical and functional impairment. There are no universally accepted scales for assessment of overall clinical and functional impairment in PD and DLB. The NSD working group reviewed validated clinical outcome assessments (COAs); the selection criteria for these anchors were: (1) COAs that measured severity of impairment across three clinical domains (cognitive, other non-motor, and motor domains) and (2) widely utilized in observational and interventional studies. Hence, the MDS-UPDRS and MoCA were selected, recognizing that a wide armamentarium of other COAs could have been utilized.

The NSD-ISS incorporates the presence of clinical signs or symptoms across three clinical domains: motor, cognitive, and non-motor. While the conceptual paper outlines a wide spectrum of motor and non-motor manifestations, many of the prodromal features are nonspecific and are common in aging, including anxiety, depression, constipation, general sleep disturbances and autonomic dysfunction. Therefore, non-motor symptoms for Stage 2 are limited to the presence of hyposmia or RBD as both are specific and predictive of PD progression^17,18^. Hyposmia was defined as UPSIT %ile ≤15 adjusted for age and sex^8^ ; RBD was defined by polysomnography-confirmed diagnostic criteria or clinical diagnosis (85% vs 15%). The MDS-UPDRS-III was used to evaluate motor signs/symptoms. In addition, we have not included non-motor symptoms as an anchor for Stage 3. Subthreshold parkinsonism, an anchor for Stage 2, was defined as MDS-UPDRS-III score ≥5, excluding the postural and action tremor items. This MDS-UPDRS-III cut-off was the mean plus two standard deviation among all PPMI healthy controls. Cognitive impairment for stage 2 was defined by MoCA total score <24 and MDS-UPDRS 1.1=1.

For the assessment of functional impairment, MDS-UPDRS-I total score and MDS-UPDRS-II total score were used to assess the severity of non-motor and motor functional impairment, respectively as these scales include items that assess instrumental and basic activities of daily living. Item 1.1 from MDS-UPDRS-I was used to assess cognitive functional impairment. The stage-based cut offs for MDS-UPDRS-I and II were selected iteratively by the NSD working group favoring a pragmatic approach. To obtain stage-specific cut-offs, we determined the upper limit for a stage by multiplying the number of items in each part (13 items) by the item severity score (i.e., slight=1, mild=2, moderate=3, severe =4). For example, a range of 14-26 was determined for Stage 4 to reflect clinical symptoms with mild functional impairment (calculation: 13 items multiplied by 2).

Based on the anchors, individuals were then categorized into one of seven stages. Table 1 lists the biologic anchors and the stage-specific cut-offs for clinical and functional impairment anchors. Importantly, only S+ individuals qualify for staging (aside from SNCA carriers who are stage 0 independent of S status). Once individuals develop disease relevant clinical signs or symptoms across any of the 3 clinical domains, they advance to Stage 2. Individuals can be S+D- (Stage 2A) or S+D+ (Stage 2B) with subtle clinical signs/symptoms defined as having one or more of the following: MDS-UPRDS item 1.1 score of 1, RBD, hyposmia, subthreshold parkinsonism, or taking PD medications. For stage 3, individuals must be S+D+ with clinical signs/symptoms as Stage 2, but these clinical signs/symptoms cause slight functional impairment. For this analysis we have selected cognitive and motor domains: cognitive (defined as item 1.1 score AND MoCA ≤ 24) and motor (defined as MDS-UPDRS-II score between 3-13). Transition from Stage 2 to 3 does not incorporate total MDS-UPDRS Part I score due to lack of specificity of the non-motor domain in this early stage. Progressively increasing impairment in any of the three domains, as measured by item 1.1, MDS-UPDRS-I total score or MDS-UPDRS-II total score, delineates Stages 4 through 6: mild (Stage 4), moderate (Stage 5), and severe (Stage 6) (Table 1).

**Table 1.**
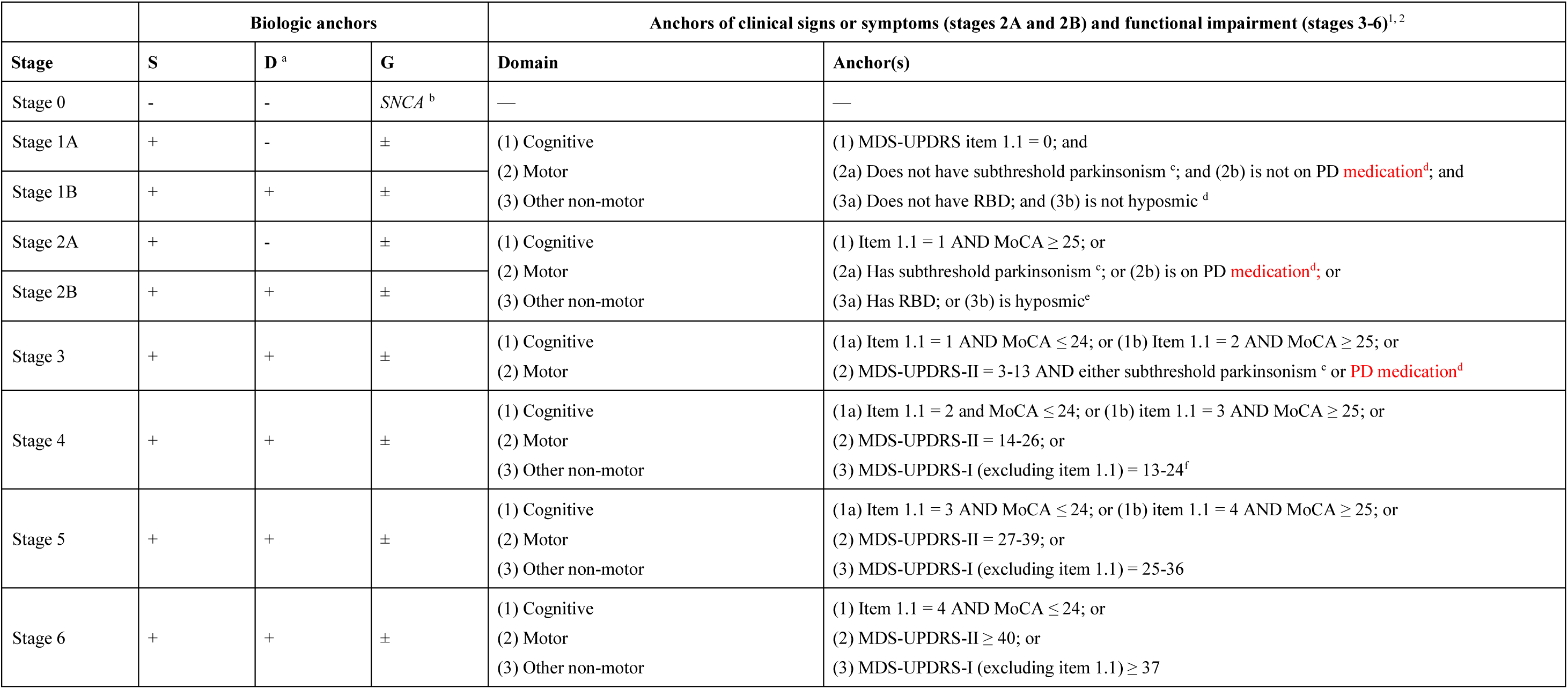
Staging anchors for application of the NSD-ISS.

Individual-level participant data available for each study are summarized in Figure 1. Participants without CSF samples for SAA testing and without an *SNCA* variant were considered not evaluable and excluded from analyses. Across the three studies, 1,741 participants with available CSF samples for αSyn-SAA testing and/or who carried an *SNCA* variant were included in analyses. Of these, 1,030 (59%) [859 PPMI, 61 PASADENA, 110 SPARK] were S+ and considered NSD, while 711 [694 PPMI, 6 PASADENA, and 11 SPARK] were S- and considered Not NSD. An additional 1, 242 participants did not have SAA results yet; thus, S status and NSD-ISS staging could not be determined.

**Figure 1.**
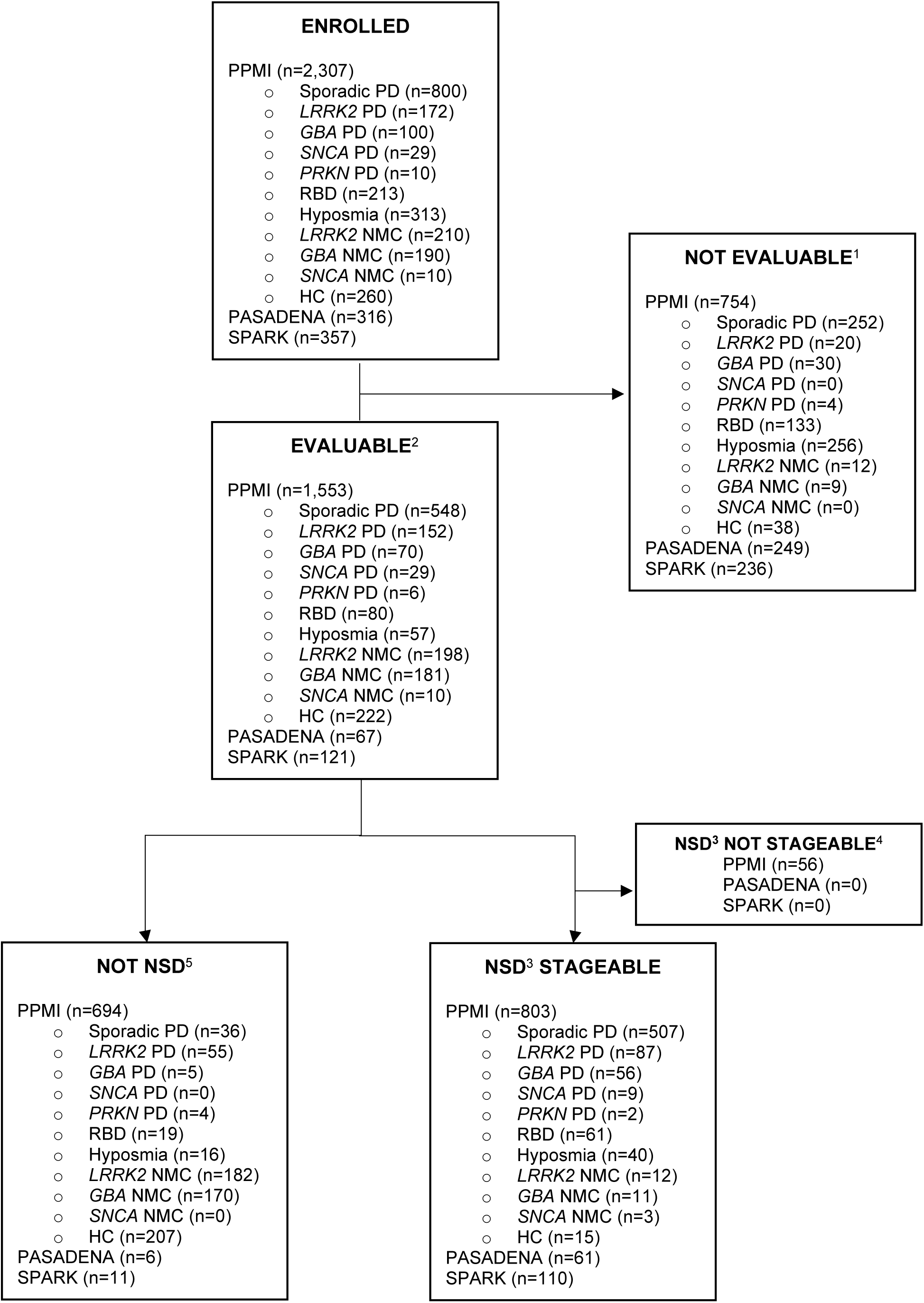
Participant Flowchart ^1^Not evaluable = CSF samples not available for alpha-synuclein aggregation testing ^2^Evaluable = CSF samples analyzed for alpha-synuclein aggregation and results available ^3^NSD = Individuals with positive alpha-synuclein aggregation tests (S+) ^4^Not stageable = Missing DaT-SPECT or clinical data and unable to assess stages ^5^Not NSD = Individuals with negative alpha-synuclein aggregation tests (S-) NSD= Neuronal Synuclein Disease

Of the PPMI participants with a clinical diagnosis of PD, 88% were S+ including 100% of *SNCA* PD, 93% of sporadic PD and *GBA* PD, 64% of *LRRK2* PD, and 33% of *PRKN* PD. Most of the S-were among healthy controls and non-manifesting genetic cohorts, including 182 *LRRK2* non-manifesting carriers and 170 *GBA* non-manifesting carriers. In PASADENA and SPARK, 91% of early PD participants with CSF samples were S+.

Table 2 shows baseline demographic characteristics of individuals with stageable NSD by study and recruitment cohort. Results were comparable between PPMI sporadic PD, PASADENA, and SPARK cohorts, considering difference in inclusion criteria. Consistent with the inclusion criteria, the genetic cohort in PPMI had longer disease duration at study entry. Baseline characteristics are also presented for Not NSD (Supplementary Table 3), Not evaluable (Supplementary Table 3a) and NSD but not stageable (Supplementary Table 3b) individuals.

**Table 2.**
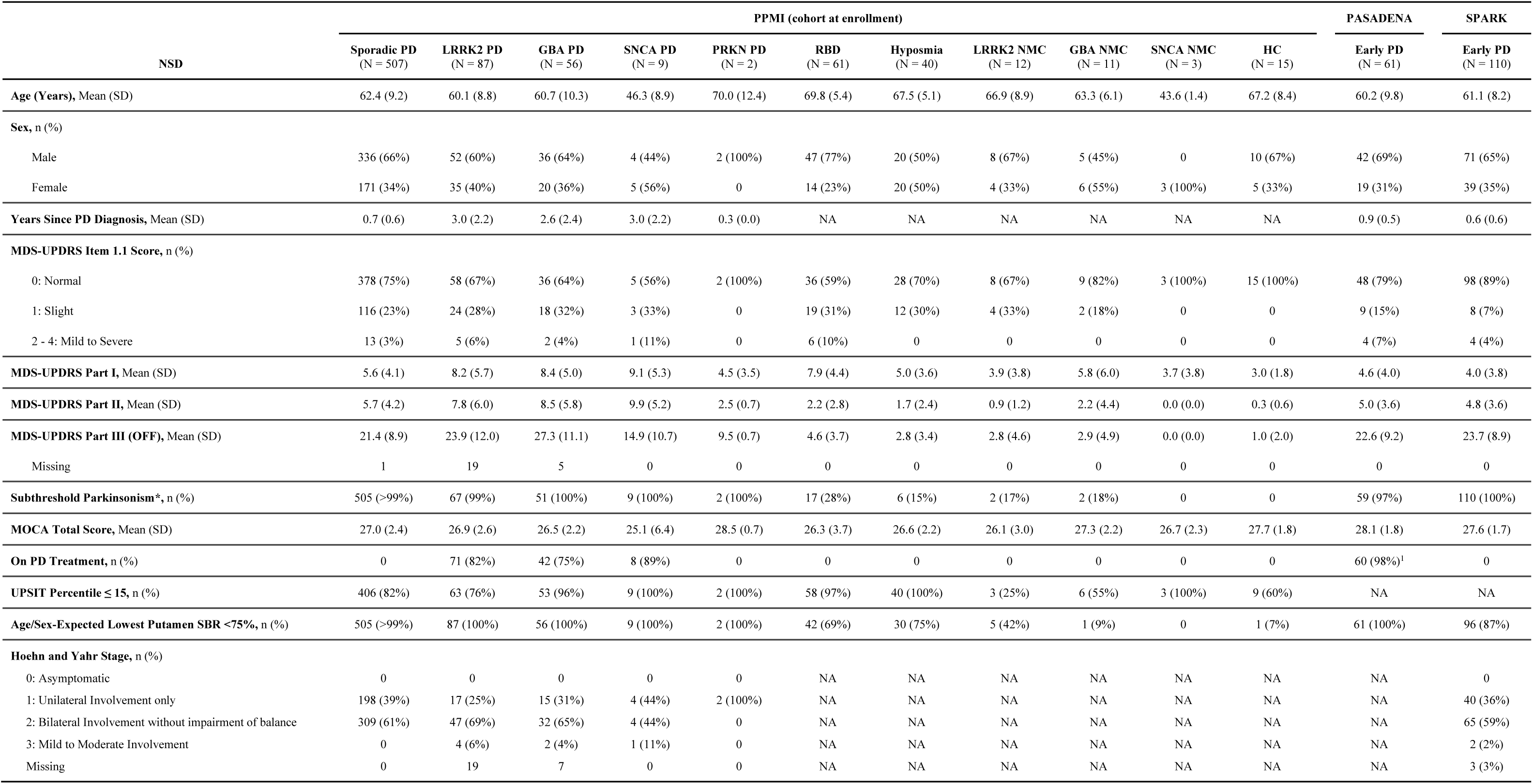
Baseline demographic characteristics of NSD participants by study.

Staging at baseline. Out of 1,030 individuals with NSD, 56 PPMI participants were missing complete data (e.g., DAT, clinical measures) and not staged. Otherwise, the prevalence of each of the NSD-ISS stages within the PPMI, PASADENA, and SPARK studies are provided in Table 3. In PPMI, most individuals with clinically diagnosed early PD met criteria for stage 3 (65% sporadic PD, 61% *LRRK2* PD, and 59% *GBA* PD); similarly, 66% PASADENA and 55% SPARK participants were stage 3. Stage 2B was the second most prevalent stage with 25% PPMI sporadic PD, 26% PASADENA, and 25% SPARK. Additionally, 13% met stage 2A criteria in the SPARK trial (Supplementary Figure 1). Across the PPMI RBD and hyposmic cohorts combined, most were stage 2A or 2B, 9% were stage 3, and 8% were stage 4 or 5. The number of non-manifesting carriers who had NSD across the genetic variants was very small (Figure 1) and the majority were in stage 1 and 2A (Table 3). Comparison of NSD-ISS by H&Y stage are presented in Table 4.

**Table 3.**
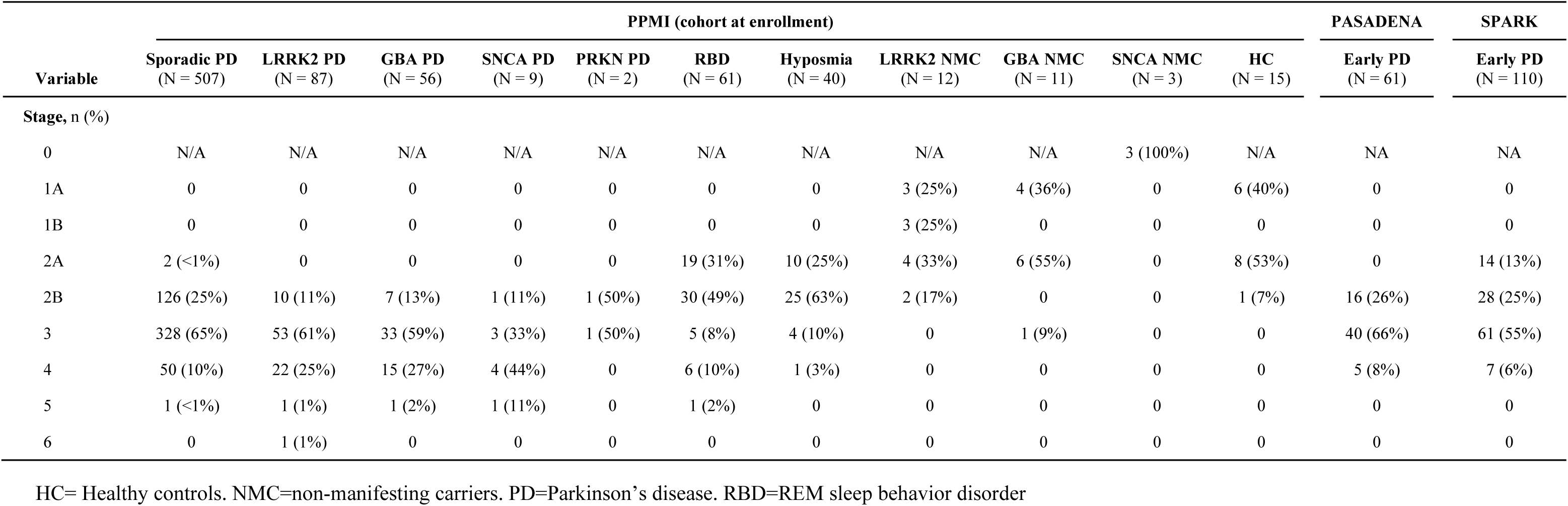
Baseline staging of NSD participants in the PPMI, PASADENA, and SPARK studies.

**Table 4.** NSD stage by Hoehn and Yahr stage among PPMI participants with PD phenotype.

|  | Hoehn and Yahr (OFF) stage* |  |  |
| --- | --- | --- | --- |
| Variable | 1<br>(N = 236) | 2<br>(N = 392) | 3<br>(N = 7) |
| NSD stage, n (%) |  |  |  |
| 0 | 0 | 0 | 0 |
| 1A | 0 | 0 | 0 |
| 1B | 0 | 0 | 0 |
| 2A | 0 | 2 (1%) | 0 |
| 2B | 81 (34%) | 62 (16%) | 0 |
| 3 | 137 (58%) | 261 (67%) | 2 (29%) |
| 4 | 16 (7%) | 64 (16%) | 5 (71%) |
| 5 | 2 (1%) | 2 (1%) | 0 |
| 6 | 0 | 1 (<1%) | 0 |

\* Hoehn & Yahr (OFF) stage was not collected at baseline for 26 participants.

To assess the impact of baseline staging on disease progression, we utilized a progression milestones approach described recently^19^. In brief, twenty-five key clinical outcomes, hereto forth termed progression milestones, spanning six clinical domains, including “walking and balance”; “motor complications”; “cognition”; “autonomic dysfunction”; “functional dependence”; and “activities of daily living”, were examined. Milestones were chosen by a working group of clinical experts, based on knowledge of the existing literature and clinical experience, and intended to reflect an unambiguously clinically meaningful and functionally relevant degree of dysfunction (e.g., postural instability, motor fluctuations, cognitive impairment, urinary incontinence, loss of functional independence, choking). A composite binary endpoint, defined as time to first occurrence of any one of the milestones, was used to assess progression.

Among 504 PPMI sporadic PD participants in stages 2B-4, 37 (7%) who met milestone criteria at baseline (5 stage 2B [4%], 20 stage 3 [6%], 12 stage 4 [24%]) and 35 (7%) without follow-up data (9 stage 2B [7%], 24 stage 3 [7%], 2 stage 4 [4%]) were excluded from the analysis. Otherwise, during a median (IQR) clinical follow-up of 7.1 (2.0, 10.1), 248/432 participants (57%) experienced a new milestone (53/112 stage 2B [47%], 168/284 stage 3 [59%], 27/36 stage 4 [75%]). The remaining participants were censored due to not reaching a milestone during the follow-up period, completing participation in the study, or loss to follow-up. Individuals in stage 4 progressed fastest, with a median (95% CI) time to developing a new milestone of 2.4 (1.0, 4.0) years compared to 5.9 (4.1, 6.0) years and 8.3 (6.2, 10.1) years among individuals in stages 3 and 2B, respectively. The survival curves differed significantly across stage strata (Χ^2^=43.6, P<.0001; Figure 2).

**Figure 2.**
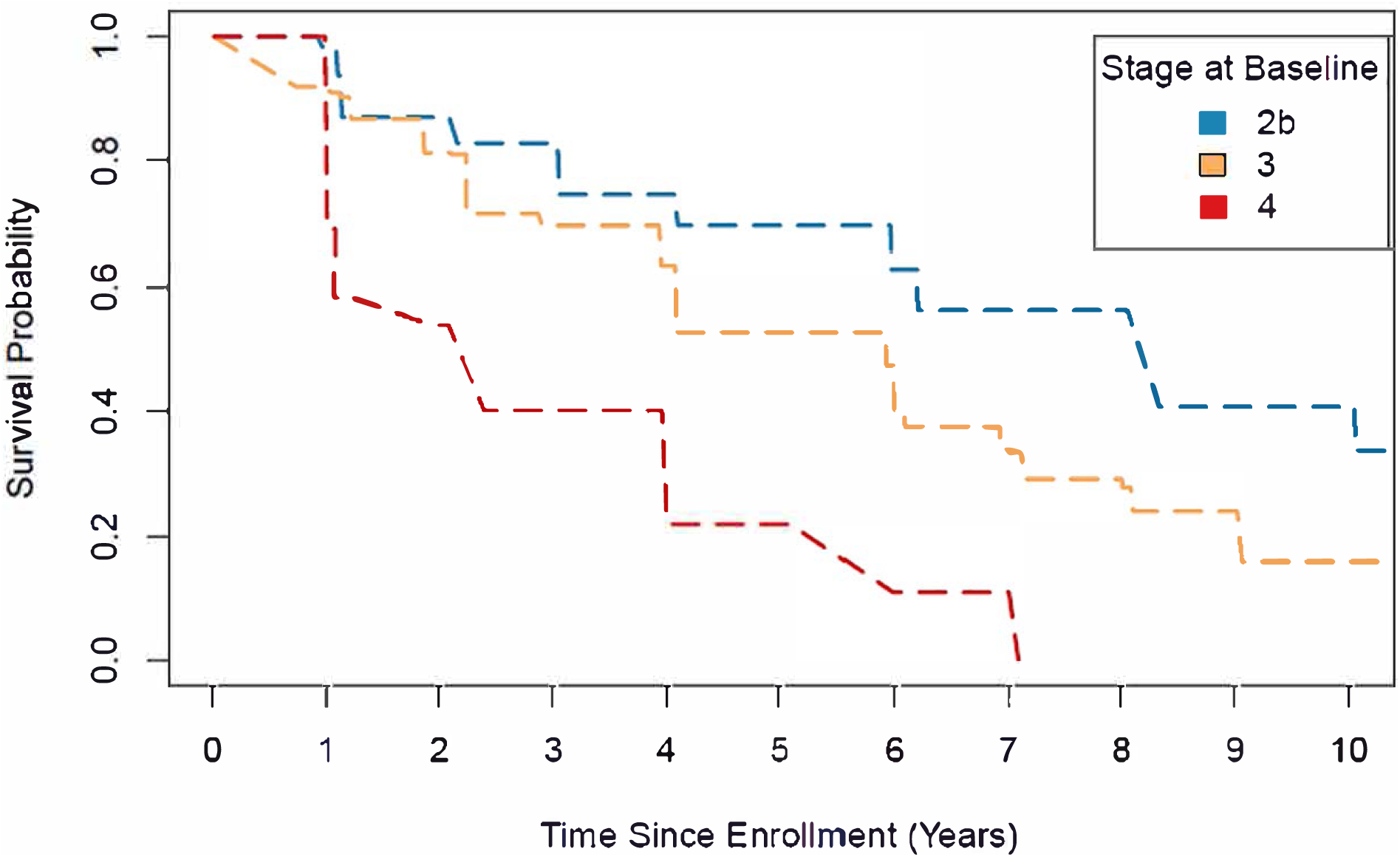
Time to reaching any progression milestone by stage at baseline among PPMI sporadic PD cohort Interval-censored survival curves of progression-free survival stratified by stage at baseline among PPMI sporadic PD participants. Progression was defined as reaching any clinically meaningful milestone across any of six clinical domains (walking and balance, motor complications, cognition, autonomic dysfunction, functional dependence, activities of daily living). Median (95% CI) progression-free survival equaled 8.3 (6.2, 10.1) years, 5.9 (4.1, 6.0) years, and 2.4 (1.0, 4.0) years among participants with a baseline stage of 2B, 3, and 4, respectively. A generalized log-rank test indicated a significant difference across the survival curves (P<0.0001).

## DISCUSSION

We position our anchors as demonstration of proof of principle and expect that the framework and operational definitions will evolve over time as new data become available. This paper is the first attempt to define biologic and clinical anchors for NSD and apply stage-specific anchors of functional impairment to well-characterized observational (PPMI) and clinical trials (PASADENA and SPARK) comprised of a broad spectrum of individuals identified as early PD, prodromal PD, non-manifesting carriers of genetic variants associated with PD, and healthy controls. Our data supports the biologic definition of NSD and highlights several observations with direct relevance to therapeutic development. Currently the field has been using clinically defined diagnostic criteria of PD or DLB and there are no established frameworks of biological and clinical subtyping. We deployed a pragmatic approach using existing scales to develop measures of worsening functional impairment within the biologically defined population, (NSD), and established a standardized staging system supported by data from three studies to operationalize the NSD-ISS conceptual framework. The thresholds were selected based on *a priori* distinction between the stages driven by severity of functional impairment (mild, moderate, severe) which are clinically meaningful and aligns with regulatory terminology. Our analyses suggest that NSD is widely applicable and present in most individuals with phenotypicPD. Across the clinical PD spectrum, approximately 90% of individuals met criteria for NSD.

While our results will need to be validated in other cohorts, they support both the face validity of the NSD-ISS and highlight the clinical heterogeneity of individuals currently identified as newly diagnosed PD;Approximately 63% of early PD individuals met criteria for Stage 3 (slight functional impairment) at baseline, consistent with expectations given the selected anchors for the stage. These individuals are the target for many clinical studies enrolling recently diagnosed untreated PD. However, on average 25% of individuals enrolled as early PD were stage 2B (with no functional impairment) and 9% were Stage 4 (with mild function impairment). These findings highlight the substantial heterogeneity in clinical and functional impairment among individuals currently considered to be early PD based on the clinical diagnosis. By defining a study population by its biology and level of functional impairment, NSD-ISS provides a paradigm that is reproducible and reduces heterogeneity, a key goal for therapeutic development. Staging with NSD-ISS better differentiates severity and is more dynamic than the widely-used H&Ystaging system. Individuals identified as H&Y stages 1 and 2 are distributed across NSD-ISS stages 2B to 4, capturing a wide range of functional impairment.

Further evidence of the face validity of the NSD-ISS is the observation that baseline stage predicts time to progression to a clinically meaningful milestone. Identifying clearly defined and reliable baseline predictors of disease progression is a crucial unmet need for PD clinical research. Individuals in Stage 4 progressed faster than Stage 2B; median time to developing a new milestone was 2.4 years versus 8.3 years,suggesting that combined biologically and functionally based staging may identify groups with less variance in progression and more power to detect change in future therapeutic trials. More data are necessary to support predictive validity of NSD-ISS for progression in early stages, especially from stages 1 to 2.

Our findings highlight the artificial nature of the current separation between prodromal disease versus early PD. Approximately 25% of the sporadic PD in PPMI and 38% of early PD in SPARK met criteria for Stage 2. Currently there is an arbitrary line between prodromal syndrome and newly diagnosed PD/DLB even though these individuals share common molecular pathologic features and similar degree of functional impairment, despite a spectrum of clinical syndromes. Our hope is that terms like “prodromal” or “early PD” will be replaced with “NSD Stage X”, that describes both the biologic underpinning and subsequent clinical and functional impairments. The NSD-ISS thus provides a framework that enables a standardized lexicon for inclusion criteria and study design, reliable findings and interpretation of results across studies. Validation in interventional studies will be essentially important.

As therapeutic development moves into the earlier stages, NSD-ISS provides a framework to identify and enrich individuals with Stage 2 for earlier interventions--spanning early motor/non-motor manifestations and individuals currently labeled as “prodromal”. The NSD-ISS enables clinical trials in individuals prior to the onset of the any of clinical manifestations that currently define PD and DLB. Notably, 76% and 72% of individuals with RBD and hyposmia were S+. As such, these S+ individuals may now be eligible for future αSyn targeted therapies. These numbers may be higher compared to other studies as the PPMI prodromal cohort was enriched with individuals with positive DAT imaging. Not surprisingly, the prevalence of NSD was lower in at risk, asymptomatic individuals with genetic variants (8% of *LRRK2* NMC, 6% of *GBA* NMC) who may not develop or have delayed development of NSD. Ultimately, interventions in Stage 0 or 1, prior to onset of any symptoms, will offer ability to test primary disease prevention strategies. In order to achive that goal, more data on the timelines of progression in early stages and baseline predictors of progression are necessary.

Despite the categorical nature of current biomarkers, this still represents substantial progress compared with a strict clinical definition of disease. There are a number of study limitations that reflect current gaps in knowledge and should guide future research. Ascertainment of S+ status is required to assess NSD criteria. We acknowledge important feasibility and scalability limitations of the CSF matrixes for n-asyn testing. We anticipate that it will transition from CSF to more accessible tissues or fluids (e.g., skin, blood) in the near future. In the interim, while not a substitute for biological characterization, a readily accessible assessment of hyposmia may significantly reduce the number of S-individuals in these trials. The NSD-ISS delineates Stage 1A versus 1B and Stage 2A versus 2B based on the hypothesis that neuronal synuclein aggregation precedes dopamine system dysfunction. Additional datasets and longitudinal follow-up of prodromal and non-manifesting genetic cohorts including ethnically diverse populations are necessary to further inform the temporal relationship between S and D positivity, and whether progression through the stages is sequential. Separation between Stage 2A and 2B in our framework is based on the selection of DAT imaging quantitative cut-off. Future studies may reexamine the cutoff which may impact the distribution of participants in Stage 2. Nevertheless, our results suggest that it is possible to enroll individuals into a clinical trial who are S+D- with subtle clinical signs/symptoms and no functional impairment. More data are necessary to define the timeline of progression from S+D- (Stage 1A) to S+D+ (Stage 1B) and from Stage 1 to 2. Future technological advances will enable quantitative biomarkers to assess progression through all stages. Neverthelesss, the framework enables targeted research in Stage 0 or 1 individuals to elucidate individual biomarker time course and inter-relationship between biomarkers essential for future disease prevention studies.

The analyzed datasets primarily enrolled individuals with motor phenotype of NSD, and data from individuals with dominant cognitive phenotype, prodromal-DLB, or DLB were limited. While there are several large phenotypically and partially biologically characterized cohorts, currently very few DLB cohorts have the biomarkers assessments required for application of the NSD-ISS. This has motivated new initiatives and work is in progress to apply NSD-ISS to several DLB consortiums and datasets.

We encourage the field to work collaboratively to explore and develop alternative functional anchors in a joint effort to advance the field towards successful therapeutic development to treat this devastating disease. Some potential areas for future iterations include evaluation of additional anchors, including novel and advanced disease-specific markers, non-motor symptoms, and functional anchors. Better delineation of the spectrum of the clinical features that signify stage 2 will inform the field. Transition to stage 3 is currently defined by functional impairment in either cognitive or motor domains; a path for the non-motor domain was not included due to lack of specificity of the symptoms and confounding due to comorbid diseases and aging. More data to define such a path will be necessary. Future NSD-ISS iterations may consider operationalizing additional non-motor signs/symptoms such as constipation, dysautonomia, and disease-related depression and anxiety, potentially drawing from other MDS-UPDRS-I items. Our selection of the anchors for Stage 3-6 was pragmatic and limited by the available measures from included studies. Additional operational definitions and analyses may be considered, such as alternative available clinical and functional scales (i.e., PDQ-39, MDS-Non-Motor Symptoms Scale, or Schwab and England Activities of Daily Living) and/or refinement of cut-offs to delineate stages. We envision that the field will require development of novel patient-centered sensitive measures of functional impairment.

In conclusion, we provide the first data-informed application of the NSD definition and the NSD-ISS. Our data strongly support the concept of the biological definition and staging framework both to optimize a study population prior to symptoms and to identify a study population with more homogenous functional impairment and disease progression at the start of symptoms. The conceptual framework and operational definitions provide an opportunity to build on the current NSD-ISS framework to further inform therapeutic development.

## METHODS

Descriptive statistics at baseline, including mean and standard deviation (SD) for continuous measures and frequency (percentage) for categorical measures, were calculated by cohort and subgroup. Results were reported separately for NSD participants who could be staged, NSD participants who could not be staged, participants without NSD, and participants not evaluable for NSD. Among NSD participants who could be staged, baseline stage was tabulated by cohort and subgroup. To assess the relationship between baseline stage and clinical progression among PPMI sporadic PD participants in stages 2B through 4, nonparametric survival function plots using the EMICM algorithm with imputed standard errors for interval-censored data were generated for time from study enrollment to reaching any progression milestone, stratified by stage. Differences in survival across stage strata were assessed using a two-sided generalized log-rank test at an alpha level of 0.05. Pointwise confidence intervals for the median time to reaching a progression milestone were obtained using a log-log transformation. Participants who did not reach a milestone or were lost to follow-up were right-censored. Participants were excluded from the analysis if they met milestone criteria at baseline and/or never completed any follow-up visits. Time to censoring and duration of follow-up were calculated as the number of years from the date of enrollment to last follow-up date. The analysis datasets comprised convenience samples, limited by available data, and formal sample size justification was not performed. Figures were created using RStudio (Posit Software, PBC, Boston, MA; posit.co; RRID:SCR 000432). All other analyses were performed using SAS v9.4 (SAS Institute Inc., Cary, NC; sas.com; RRID:SCR 008567).

## Supporting information

PPMI Study Group Authors

## ACKNOWLEDGEMENTS

We appreciate Dr. Teresa Buraccio for critical review of the draft of the manuscript.

We thank the SPARK investigators, trial coordinators, staff, and participants for their contribution to the trial. We appreciate Dr. Minhua Yang for analyses of the SPARK data.

We thank the PASADENA investigators, trial coordinators, staff, and participants for their contribution to the trial. We appreciate Drs. Thomas Riorge and Annabelle Monnet for analyses of the PASADENA data.

## Sources of funding

Funding support for the data analysis was provided by The Michael J. Fox Foundation for Parkinson’s Research (MJFF). This report presents data from three studies, including PASADENA, supported by F. Hoffman-La Roche and Prothena Biosciences, SPARK, supported by Biogen, and PPMI. PPMI – a public-private partnership – is funded by MJFF and funding partners, including 4D Pharma, Abbvie, AcureX, Allergan, Amathus Therapeutics, Aligning Science Across Parkinson’s, AskBio, Avid Radiopharmaceuticals, BIAL, BioArctic, Biogen, Biohaven, BioLegend, BlueRock Therapeutics, BristolMyers Squibb, Calico Labs, Capsida Biotherapeutics, Celgene, Cerevel Therapeutics, Coave Therapeutics, DaCapo Brainscience, Denali, Edmond J. Safra Foundation, Eli Lilly, Gain Therapeutics, GE HealthCare, Genentech, GSK, Golub Capital, Handl Therapeutics, Insitro, Janssen Neuroscience, Jazz Pharmaceuticals, Lundbeck, Merck, Meso Scale Discovery, Mission Therapeutics, Neurocrine Biosciences, Neuropore, Pfizer, Piramal, Prevail Therapeutics, Roche, Sanofi, Servier, Sun Pharma Advanced Research Company, Takeda, Teva, UCB, Vanqua Bio, Verily, Voyager Therapeutics, the Weston Family Foundation and Yumanity Therapeutics.

## Role of the funding source

Research officers (SC, MF, CK, TS, YX) at MJFF, PPMI Sponsor, were involved in the design of the study and writing of the manuscript.

## Data source

PPMI data used in the preparation of this article were obtained June 12, 2023 from the Parkinson’s Progression Markers Initiative (PPMI) database (www.ppmi-info.org/access-data-specimens/download-data), RRID:SCR 006431. For up-to-date information on the study, visit www.ppmi-info.org.

## Data availability statement

PPMI data are publicly available from the Parkinson’s Progression Markers Initiative (PPMI) database (www.ppmi-info.org/access-data-specimens/download-data), RRID:SCR 006431. For up-to-date information on the study, visit www.ppmi-info.org. Use of Tier 4 data: This analysis was conducted by the PPMI Statistics Core and used actual dates of activity for participants, a restricted data element not available to public users of PPMI data

For PASADENA, qualified researchers may request access to individual patient level clinical data through a data request platform. At the time of writing, this request platform is Vivli. https://vivli.org/ourmember/roche/. Further details on Roche’s criteria for eligible trials for data access are available here (https://vivli.org/members/ourmembers/). For up-to-date details on Roche’s Global Policy on the Sharing of Clinical Information and how to request access to related clinical study documents, see here: https://go.roche.com/data_sharing. Anonymized records for individual patients across more than one data source external to Roche cannot, and should not, be linked due to a potential increase in risk of patient re-identification.

For SPARK, to request access to data, please visits http://www.biogenclinicaldatarequest.com. The individual participant data collected during the trial, which supports the research proposal, will be available to qualified researchers after anonymization and upon approval of the research proposal. Anonymization of the datasets is necessary to allow data to be shared ethically and legally, and to maximize their significant social, environmental, and economic value, whilst preserving confidentiality of the individuals who participated in studies conducted by Biogen.

## CODE AVAILABILITY STATEMENT

Not applicable as per the journal guidelines.

## COMPETING INTERESTS

TD declares former employment for and employee stock options in Biogen. GP declares employment for F. Hoffmann-La Roche Ltd. and stock ownership for F. Hoffmann-La Roche Ltd., Atea, Novartis and Eli Lilly. MB declares travel grants from The Michael J. Fox Foundation. CG declares employment for The Michael J. Fox Foundation. KP declares consultancies for Curasen; was on a scientific advisory board for Curasen and Amprion; honoraria from invited scientific presentations to universities and professional societies not exceeding $5000 per year from California Congress of Clinical Neurology, California Neurological Society, and Johns Hopkins University; and patents or patent applications numbers 17/314,979 and 63/377,293. KP also declares grants to her institution (Stanford University School of Medicine) from NIH/NINDS NS115114, NS062684, NS075097, NIH/NIA U19 AG065156, P30 AG066515, The Michael J. Fox Foundation, Lewy Body Dementia Association, Alzheimer’s Drug Discovery Foundation, Sue Berghoff LBD Research Fellowship and the Knight Initiative for Brain Resilience. DW declares salary support from The Michael J. Fox Foundation for serving on an Executive Steering Committee for the PPMI and consultancies for Roche Pharma. DW declares membership on the npj Parkinson’s Disease Editorial Board. LC declares grants to her institution from Biogen (clinical trial funding), MJFF, UPMC Competitive Medical Research Fund, National Institutes of Health, and University of Pittsburgh; grant and travel support from MJFF; royalties from Wolters Kluwel (for authorship); and in-kind donation by Advanced Brain Monitoring of equipment for research study to her institution. CC declares grants from The Michael J. Fox Foundation and NIH/NINDS. CT declares consultancies for CNS Ratings, Australian Parkinson’s Mission, Biogen, Evidera, Cadent (data safety monitoring board), Adamas (steering committee), Biogen (via the Parkinson Study Group steering committee), Kyowa Kirin (advisory board), Lundbeck (advisory board), Jazz/Cavion (steering committee), Acorda (advisory board), Bial (DMC) and Genentech. CT also declares grant support to her institution from The Michael J. Fox Foundation, National Institute of Health, Gateway LLC, Department of Defense, Roche Genentech, Biogen, Parkinson Foundation and Marcus Program in Precision Medicine. CT declares membership on the npj Parkinson’s Disease Editorial Board. CK declares employment for The Michael J. Fox Foundation. YX declares employment for and travel grants from The Michael J. Fox Foundation. SC declares employment for and travel grants from The Michael J. Fox Foundation. LC-M declares employment for and employee stock options in Amprion, grants 16712 and 21233 from The Michael J. Fox Foundation to his institution; grant U44NS111672 from NIH to his institution; and patents or patent application numbers US 20210277076A1, US 20210311077A1, US 20190353669A1 and US 20210223268A1. PD has no declarations. TF declares travel grants and grant payments to her institution (Indiana University) from The Michael J. Fox Foundation. MF declares employment for The Michael J. Fox Foundation and an unpaid advisory role at Vaxxinity. DJ declares employment for and employee stock options from Denali Therapeutics. KK declares support to his institution (University of Rochester Medical Center) from The Michael J. Fox Foundation. KMe declares consultancies for The Michael J. Fox Foundation, AcuRx, Caraway, Cerebral Therapeutics, NRG Therapeutics (scientific advisory board), Nitrase Therapeutics (scientific advisory board), Nurabio, Retromer Therapeutics (director on the board, part-time chief scientific officer), Schrodinger, Sinopia Biosciences (scientific advisory board), and Vanqua Biosciences (scientific advisory board); stock ownership for Cognition Therapeutics, Eli Lilly (retiree stock holder), Envisagenics, Nitrase Therapeutics, Sinopia Biosciences and Retromer Therapeutics; honoraria for the University of Utah; patents or patent applications for Retromer Therapeutics (planned patent); research grant from The Michael J. Fox Foundation and travel grants from the University of Utah. BM declares consultancies from Roche and Biogen; grants from The Michael J. Fox Foundation, ASAP and DFG; honoraria for Abbvie and Bial; leadership role for The Michael J. Fox Foundation and travel grants for Abbvie. TM declares support to his institution (Stanford University School of Medicine) from The Michael J. Fox Foundation. KN declares grant to her institution from The Michael J. Fox Foundation. JS declares consultancies from Invicro, Biogen, and Abbvie; and stock ownership from RealmIDX, MNI Holdings, and LikeMinds as well as grants from The Michael J. Fox Foundation. TSh declares employment for The Michael J. Fox Foundation. ASin declares employment for The National Institutes of Health who received grants from The Michael J. Fox Foundation and ASAP. ASin declares Diagnostic for Stroke royalties (unrelated to current work); honoraria from Movement Disorders Society and Nature Publishing Group; travel grants from Chan Zuckerberg Initiative, The Michael J. Fox Foundation and Weill Cornell. ASin’s spouse is an employee of GeneDx. ASin declares being an associate editor for the npj Parkinson’s Disease Editorial Board. DS has no declarations. MS declared consultancies for Mediflix, Inc., Health and Wellness Partners; honoraria from Atria Foundation, International Parkinson and Movement Disorder Society, Neurocrine, Luye Pharma and Acorda. MS serves on advisory board at Neuroderm, Alexza, Alexion and Biogen. CS declares employment for Amprion; stock ownership for Amprion; honoraria (will receive royalties for the sale of seed amplification assay [SAA]) from Amprion; and patents or patent applications, awarded and amplified in conjunction with Amprion for the SAA assay. ET has no declarations. ASid declares consultancies for SPARC Therapeutics, Capsida Therapeutics and Parkinson Study Group; honoraria from Bial; grants from The Michael J. Fox Foundation (member of PPMI Steering Committee); and participation on board at Wave Life Sciences, Inhibikase, Prevail, Huntington Study Group and Massachusetts General Hospital. BD declarations are TBD. TSi declares consultancies for AcureX, Adamas, AskBio, Amneal, Blue Rock Therapeutics, Critical Path for Parkinson’s Consortium, Denali, The Michael J. Fox Foundation, Neuroderm, Roche, Sanofi, Sinopia, Takeda, and Vanqua Bio; on advisory boards for AcureX, Adamas, AskBio, Biohaven, Denali, GAIN, Neuron23 and Roche; on scientific advisory boards for Koneksa, Neuroderm, Sanofi and UCB; and received research funding from Amneal, Biogen, Roche, Neuroderm, Sanofi, Prevail and UCB and an investigator for NINDS, MJFF, Parkinson’s Foundation. KMa declares support to his institution (Institute for Neurodegenerative Disorders) from The Michael J. Fox Foundation. KMa also declares consultancies for Invicro, The Michael J. Fox Foundation, Roche, Calico, Coave, Neuron23, Orbimed, Biohaven, Anofi, Koneksa, Merck, Lilly, Inhibikase, Neuramedy, IRLabs and Prothena and participates on DSMB at Biohaven.

## AUTHOR CONTRIBUTIONS

TD, GP, KP, DW, LC, CT, CK, YX, SC, LC-M, PD, TF, MF, DJ, KK, KM, BM, TM, KN, JS, TSh, ASin, DS, MS, CS, ET, ASi, BD, TSi, KM contributed to the conceptualization. MB, CG, LC, CC, ASid, BD, TSi and KMa contributed to the data curation, formal analysis and validation. TD, GP, CG, KP, DW, LC, CT, LC-M, PD, TF, DJ, KK, KM, BM, TM, KN, JS, ASin, DS, MS, CS, ET, ASi, BD, TSi, KMa contributed to the supervision. MB, CG, LC, CC, ASid, BD, TSi and KMa contributed to the writing of the original draft. CK, YX, SC, MF and TSh acquired funding, provided resources and contributed to the project administration. All authors contributed to the investigations, methodology and the review and editing of the final version of the manuscript. TD and TSi have had direct access to and verified all components of the submitted manuscript.

## Notes

### Summary of Updates

This version has been updated to reflect minor editorial changes.

