## Supplement Materials for "Neuronal alpha-Synuclein Disease Integrated Staging System performance in PPMI, PASADENA, and SPARK baseline cohorts"

#### SUPPLEMENTARY FIGURE AND TABLES LEGENDS

Supplementary Table 1. Proposed neuronal $\alpha$-synuclein disease integrated staging system

Supplementary Table 2. Glossary of terms

Supplementary Table 3. Baseline demographic characteristics of Not NSD participants by cohort

Supplementary Table 3a. Baseline demographic characteristics of Not Evaluable participants by cohort

Supplementary Table 3b. Baseline demographic characteristics of NSD and Not Stageable participants by cohort

Supplementary Figure 1. Proportion in each NSD stage in PPMI vs. PASADENA vs. SPARK

Supplementary Table 1.

| | | a-synuclein Biomarker (S) | Dopamine Dysfunction Biomarker (D) | Clinical signs and symptoms attributable to neuronal a-synuclein disease syndromes | Functional impairment attributable to neuronal a-synuclein disease syndromes |
|---|---|---|---|---|---|
| Genetic Risk | | | | | |
| $R^L$ | (G) Genetic risk variants – low age adjusted risk (constantly redefined). | Absent | Absent | No clinical signs or symptoms | No functional impairment |
| $R^H$ | (G) Genetic risk variants – high age adjusted risk (constantly redefined) | Absent | Absent | No clinical signs or symptoms | No functional impairment |
| Stage Definition | | | | | |
| 0 | Fully penetrant *SNCA* variant (G+) | S- | D- | No clinical signs or symptoms | No functional impairment |
| 1A | Characteristic pathological changes, but no evidence of clinical signs or symptoms | S+ | D- | No clinical signs or symptoms | No functional impairment |
| 1B | Characteristic pathologic changes plus dopaminergic dysfunction, but no evidence of clinical signs or symptoms | S+ | D+ | No clinical signs or symptoms | No functional impairment |
| 2A | Characteristic pathological changes and subtle detectable clinical signs and symptoms, but no functional impairment. | S+ | D- | Subtle clinical signs or symptoms; can be motor or non-motor: hyposmia, RBD, cognitive abnormalities, constipation, dysautonomia, depression, anxiety. | No functional impairment |
| 2B | Characteristic pathologic changes plus dopaminergic dysfunction and subtle detectable clinical signs and symptoms, but no functional impairment. | S+ | D+ | Subtle clinical signs or symptoms; can be motor or non-motor: hyposmia, RBD, cognitive abnormalities, constipation, dysautonomia, depression, anxiety. | No functional impairment |
| 3 | Characteristic pathologic changes plus dopaminergic dysfunction and clinical signs and symptoms causing slight functional impairment. | S+ | D+ | Relevant motor and non-motor signs and symptoms increasing in severity, but stage is defined by a slight degree of functional impairment | Slight: functional impairment with minimal impact on complex tasks of daily life and usual activities, such as finances, transportation, food, household, conversation |
| 4 | Characteristic pathologic changes plus dopaminergic dysfunction and clinical signs and symptoms causing mild functional impairment. | S+ | D+ | Relevant motor and non-motor signs and symptoms increasing in severity, but stage is defined by a mild degree of functional impairment | Mild: functional impairment severe enough to cause some impairment in complex tasks of daily life and usual activities, but basic tasks of daily life related to personal care are intact, such as bathing, dressing, walking, using the toilet, eating |
| 5 | Characteristic pathologic changes plus dopaminergic dysfunction and clinical signs and symptoms causing moderate functional impairment. | S+ | D+ | Relevant motor and non-motor signs and symptoms increasing in severity, but stage is defined by a moderate degree of functional impairment | Moderate: functional impairment severe enough to require assistance with basic tasks of daily life |
| 6 | Characteristic pathologic changes plus dopaminergic dysfunction and clinical signs and symptoms causing severe functional impairment | + | + | Relevant motor and non-motor signs and symptoms increasing in severity, but stage is defined by a severe degree of functional impairment | Severe: functional impairment severe enough to require dependence on others for basic tasks of daily life |

Supplementary Table 2.

**Neuronal α-synuclein (n-asyn)**
Disease-defining form of a-synuclein: misfolded, pathological, predominantly neuronal a-synuclein

**Neuronal α-synuclein disease (NSD)**
Disease defined by presence of n-asyn and dopaminergic dysfunction or presence of a fully penetrant *SNCA* variant; disease is defined independent of presence of clinical signs and symptoms

**S anchor for neuronal α-synuclein disease (S)**
Indicates presence (S+) or absence (S-) of n-asyn as measured by any validated biomarker of n-asyn pathology

**D anchor for neuronal α-synuclein disease (D)**
Indicates presence (D+ or absence (D-) of dopaminergic dysfunction as measured by any validated biomarker of dopaminergic dysfunction

**Genetic status (G)**
Indicates presence (G+) or absences (G-) of relevant pathogenic variants

**α-synuclein seed amplification assay (SAA)**
An assay that leverages the self-replicating properties of misfolded a-synuclein by means of fragmentation and elongation cycles

**Not evaluable**
CSF samples not available for n-asyn SAA

**Evaluable**
CSF samples analyzed for n-asyn SAA and results available

**Not stageable**
Missing dopamine transporter or clinical data and unable to assess stages

**Not Neuronal a-synuclein disease (Not NSD)**
Disease defined by absence of n-asyn

### Supplementary Table 3.

| | PPMI (cohort at enrollment) | | | | | | | | | | | PASADENA | SPARK |
|---|---|---|---|---|---|---|---|---|---|---|---|---|---|
| NSD | **Sporadic PD** (N = 36) | **LRRK2 PD** (N = 55) | **GBA PD** (N = 5) | **SNCA PD** (N = 0) | **PRKN PD** (N = 4) | **RBD** (N = 19) | **Hyposmia** (N = 16) | **LRRK2 NMC** (N = 182) | **GBA NMC** (N = 170) | **SNCA NMC** (N = 0) | **HC** (N = 207) | **Early PD** (N = 6) | **Early PD** (N = 11) |
| **Age (Years),** Mean (SD) | 65.7 (10.9) | 68.3 (6.7) | 62.1 (10.7) | NA | 43.8 (14.2) | 66.6 (7.1) | 68.4 (6.5) | 61.3 (7.5) | 62.0 (6.8) | NA | 61.1 (11.7) | NA | 57.6 (57.7) |
| **Sex,** n (%) | | | | | | | | | | | | | |
| Male | 21 (58%) | 20 (36%) | 2 (40%) | NA | 3 (75%) | 16 (84%) | 6 (38%) | 75 (41%) | 69 (41%) | NA | 129 (62%) | NA | 6 (55%) |
| Female | 15 (42%) | 35 (64%) | 3 (60%) | NA | 1 (25%) | 3 (16%) | 10 (63%) | 107 (59%) | 101 (59%) | NA | 78 (38%) | NA | 5 (45%) |
| **Years Since PD Diagnosis,** Mean (SD) | 0.5 (0.4) | 2.8 (2.1) | 1.7 (2.1) | NA | 6.6 (2.9) | NA | NA | NA | NA | NA | NA | NA | 1.0 (0.8) |
| **MDS-UPDRS Item 1.1 Score,** n (%) | | | | | | | | | | | | | |
| 0: Normal | 29 (81%) | 39 (71%) | 3 (60%) | NA | 4 (100%) | 12 (63%) | 12 (75%) | 157 (87%) | 137 (81%) | NA | 189 (92%) | NA | 9 (82%) |
| 1: Slight | 4 (11%) | 15 (27%) | 2 (40%) | NA | 0 | 7 (37%) | 4 (25%) | 21 (12%) | 27 (16%) | NA | 16 (8%) | NA | 2 (18%) |
| 2 - 4: Mild to Severe | 3 (8%) | 1 (2%) | 0 | NA | 0 | 0 | 0 | 3 (2%) | 6 (4%) | NA | 1 (<1%) | NA | 0 |
| Missing | 0 | 0 | 0 | NA | 0 | 0 | 0 | 1 | 0 | NA | 1 | NA | 0 |
| **MDS-UPDRS Part I,** Mean (SD) | 7.2 (5.6) | 7.3 (6.2) | 7.4 (7.1) | NA | 6.0 (3.6) | 7.2 (5.5) | 5.4 (4.6) | 4.4 (3.6) | 5.6 (4.7) | NA | 3.0 (2.9) | NA | 4.3 (3.3) |
| Missing | 0 | 0 | 0 | NA | 0 | 0 | 0 | 1 | 0 | NA | 1 | NA | 0 |
| **MDS-UPDRS Part II,** Mean (SD) | 7.4 (4.7) | 5.7 (4.9) | 6.8 (4.4) | NA | 5.0 (3.2) | 1.7 (2.3) | 1.4 (1.5) | 1.0 (1.9) | 1.2 (2.2) | NA | 0.4 (0.9) | NA | 4.7 (3.9) |
| Missing | 0 | 0 | 0 | NA | 0 | 0 | 0 | 1 | 0 | NA | 1 | NA | 0 |
| **MDS-UPDRS Part III (OFF),** Mean (SD) | 22.8 (7.1) | 18.1 (8.0) | 24.8 (10.9) | NA | 16.0 (NA) | 5.4 (3.9) | 4.2 (4.0) | 3.0 (3.9) | 2.4 (3.5) | NA | 1.3 (2.2) | NA | 22.1 (9.3) |
| Missing | 0 | 9 | 1 | NA | 3 | 1 | 1 | 3 | 1 | NA | 3 | NA | 0 |
| **Subthreshold Parkinsonism*,** n (%) | 36 (100%) | 45 (98%) | 4 (100%) | NA | 1 (100%) | 8 (44%) | 7 (47%) | 29 (16%) | 23 (14%) | NA | 12 (6%) | NA | 11 (100%) |
| Missing | 0 | 9 | 1 | NA | 3 | 1 | 1 | 3 | 1 | NA | 3 | NA | 0 |
| **MOCA Total Score,** Mean (SD) | 26.9 (2.1) | 25.2 (3.2) | 27.0 (2.0) | NA | 27.5 (2.4) | 26.6 (3.1) | 27.0 (1.9) | 26.8 (2.5) | 26.7 (2.3) | NA | 28.0 (1.5) | NA | 26.3 (2.1) |
| Missing | 0 | 0 | 0 | NA | 0 | 0 | 0 | 1 | 0 | NA | 0 | NA | 0 |
| **On PD Treatment,** n (%) | 0 | 36 (65%) | 2 (40%) | NA | 0 | 0 | 0 | 0 | 1 (1%) | NA | 0 | NA | 0 |
| Missing | 0 | 0 | 0 | NA | 0 | 1 | 0 | 3 | 0 | NA | 3 | NA | 0 |
| **UPSIT Percentile ≤ 15,** n (%) | 5 (15%) | 13 (25%) | 0 | NA | 2 (50%) | 4 (21%) | 15 (94%) | 24 (13%) | 15 (9%) | NA | 21 (10%) | NA | NA |
| Missing | 2 | 2 | 0 | NA | 0 | 0 | 0 | 1 | 0 | NA | 2 | NA | NA |
| **Age/Sex-Expected Lowest Putamen SBR <75%,** n (%) | 32 (91%) | 48 (98%) | 5 (100%) | NA | 4 (100%) | 8 (42%) | 6 (38%) | 30 (17%) | 8 (5%) | NA | 28 (14%) | NA | 6 (55%) |
| Missing | 1 | 6 | 0 | NA | 0 | 0 | 0 | 5 | 5 | NA | 3 | NA | 0 |

| NSD | PPMI (cohort at enrollment) | | | | | | | | | | | PASADENA | SPARK |
|---|---|---|---|---|---|---|---|---|---|---|---|---|---|
| | Sporadic PD (N = 36) | LRRK2 PD (N = 55) | GBA PD (N = 5) | SNCA PD (N = 0) | PRKN PD (N = 4) | RBD (N = 19) | Hyposmia (N = 16) | LRRK2 NMC (N = 182) | GBA NMC (N = 170) | SNCA NMC (N = 0) | HC (N = 207) | Early PD (N = 6) | Early PD (N = 11) |
| **Hoehn and Yahr Stage,** n (%) | | | | | | | | | | | | | |
| 0: Asymptomatic | 0 | 0 | 0 | NA | 0 | NA | NA | NA | NA | NA | NA | NA | 0 |
| 1: Unilateral Involvement only | 5 (14%) | 12 (26%) | 1 (25%) | NA | 0 | NA | NA | NA | NA | NA | NA | NA | 2 (18%) |
| 2: Bilateral Involvement without impairment of balance | 31 (86%) | 29 (63%) | 3 (75%) | NA | 1 (100%) | NA | NA | NA | NA | NA | NA | NA | 9 (82%) |
| 3: Mild to moderate involvement | 0 | 5 (11%) | 0 | NA | 0 | NA | NA | NA | NA | NA | NA | NA | 0 |
| Missing | 0 | 9 | 1 | NA | 3 | NA | NA | NA | NA | NA | NA | NA | 0 |

* MDS-UPDRS Part III > 4 excluding postural and action tremor.

HC= Healthy controls. MDS-UPDRS=Movement Disorder Society Unified Parkinson's Disease Rating Scale. MoCA=Montreal Cognitive Assessment. NMC=non-manifesting carriers. PD=Parkinson's disease. RBD=REM sleep behavior disorder. SBR= Striatal Binding Ratio. UPSIT= University of Pennsylvania Smell Identification Test

### Supplementary Table 3a.

| | PPMI (cohort at enrollment) | | | | | | | | | | | PASADENA | SPARK |
|---|---|---|---|---|---|---|---|---|---|---|---|---|---|
| **NSD** | **Sporadic PD** (N = 252) | **LRRK2 PD** (N = 20) | **GBA PD** (N = 30) | **SNCA PD** (N = 0) | **PRKN PD** (N = 4) | **RBD** (N = 133) | **Hyposmia** (N = 256) | **LRRK2 NMC** (N = 12) | **GBA NMC** (N = 9) | **SNCA NMC** (N = 0) | **HC** (N = 38) | **Early PD** (N = 249) | **Early PD** (N = 236) |
| **Age (Years),** Mean (SD) | 64.2 (9.5) | 68.4 (10.3) | 62.3 (11.6) | NA | 51.5 (10.9) | 68.4 (5.4) | 67.6 (5.7) | 63.2 (7.7) | 58.9 (8.8) | NA | 61.0 (11.3) | NA | 59.7 (9.4) |
| Missing | 0 | 0 | 0 | NA | 0 | 0 | 1 | 0 | 0 | NA | 0 | NA | 0 |
| **Sex,** n (%) | | | | | | | | | | | | | |
| Male | 164 (65%) | 5 (25%) | 12 (40%) | NA | 3 (75%) | 107 (80%) | 101 (40%) | 4 (33%) | 2 (22%) | NA | 23 (61%) | NA | 173 (73%) |
| Female | 88 (35%) | 15 (75%) | 18 (60%) | NA | 1 (25%) | 26 (20%) | 154 (60%) | 8 (67%) | 7 (78%) | NA | 15 (39%) | NA | 63 (27%) |
| Missing | 0 | 0 | 0 | NA | 0 | 0 | 1 | 0 | 0 | NA | 0 | NA | 0 |
| **Years Since PD Diagnosis,** Mean (SD) | 0.8 (0.6) | 3.3 (2.0) | 3.5 (2.2) | NA | 5.7 (5.3) | NA | NA | NA | NA | NA | NA | NA | 0.7 (0.7) |
| Missing | 1 | 0 | 0 | NA | 0 | NA | NA | NA | NA | NA | NA | NA | 0 |
| **MDS-UPDRS Item 1.1 Score,** n (%) | | | | | | | | | | | | | |
| 0: Normal | 167 (66%) | 11 (55%) | 22 (73%) | NA | 3 (75%) | 81 (62%) | 185 (73%) | 6 (50%) | 7 (78%) | NA | 34 (92%) | NA | 195 (83%) |
| 1: Slight | 71 (28%) | 7 (35%) | 4 (13%) | NA | 0 | 35 (27%) | 58 (23%) | 6 (50%) | 2 (22%) | NA | 2 (5%) | NA | 41 (17%) |
| 2 - 4: Mild to Severe | 14 (6%) | 2 (10%) | 4 (13%) | NA | 1 (25%) | 15 (11%) | 11 (4%) | 0 | 0 | NA | 1 (3%) | NA | 0 |
| Missing | 0 | 0 | 0 | NA | 0 | 2 | 2 | 0 | 0 | NA | 1 | NA | 0 |
| **MDS-UPDRS Part I,** Mean (SD) | 6.4 (4.8) | 8.8 (5.6) | 7.4 (5.0) | NA | 5.5 (4.4) | 7.6 (5.9) | 6.0 (4.6) | 7.0 (4.7) | 9.0 (9.3) | NA | 3.5 (3.2) | NA | 4.4 (3.5) |
| Missing | 6 | 0 | 0 | NA | 0 | 3 | 7 | 0 | 0 | NA | 1 | NA | 0 |
| **MDS-UPDRS Part II,** Mean (SD) | 6.9 (4.9) | 6.6 (5.8) | 7.3 (6.2) | NA | 5.3 (4.6) | 2.5 (3.5) | 2.1 (3.7) | 2.4 (4.5) | 3.4 (7.6) | NA | 0.5 (1.1) | NA | 5.6 (3.9) |
| Missing | 2 | 0 | 0 | NA | 0 | 2 | 3 | 0 | 0 | NA | 1 | NA | 0 |
| **MDS-UPDRS Part III (OFF),** Mean (SD) | 25.0 (11.5) | 23.1 (8.7) | 22.5 (8.7) | NA | 15.5 (5.3) | 4.2 (4.3) | 4.1 (4.8) | 3.3 (3.0) | 5.6 (8.8) | NA | 1.6 (1.9) | NA | 22.3 (9.0) |
| Missing | 1 | 7 | 11 | NA | 0 | 2 | 16 | 0 | 1 | NA | 2 | NA | 0 |
| **Subthreshold Parkinsonism*,** n (%) | 249 (99%) | 13 (100%) | 19 (100%) | NA | 4 (100%) | 33 (25%) | 58 (24%) | 2 (17%) | 2 (25%) | NA | 3 (8%) | NA | 235 (100%) |
| Missing | 1 | 7 | 11 | NA | 0 | 2 | 16 | 0 | 1 | NA | 2 | NA | 0 |
| **MOCA Total Score,** Mean (SD) | 26.7 (2.3) | 24.6 (4.0) | 26.3 (3.7) | NA | 25.5 (3.7) | 26.1 (2.9) | 26.9 (2.2) | 26.2 (2.7) | 26.7 (2.7) | NA | 27.8 (1.6) | NA | 27.4 (2.0) |
| Missing | 3 | 1 | 0 | NA | 0 | 1 | 5 | 0 | 0 | NA | 0 | NA | 0 |
| **On PD Treatment,** n (%) | 0 | 14 (70%) | 21 (70%) | NA | 1 (25%) | 1 (1%) | 0 | 0 | 0 | NA | 0 | NA | 0 |
| Missing | 1 | 0 | 0 | NA | 0 | 2 | 6 | 0 | 1 | NA | 1 | NA | 0 |
| **UPSIT Percentile ≤ 15,** n (%) | 215 (88%) | 8 (47%) | 19 (70%) | NA | 3 (75%) | 96 (74%) | 254 (>99%) | 3 (25%) | 2 (25%) | NA | 14 (40%) | NA | NA |
| Missing | 7 | 3 | 3 | NA | 0 | 3 | 1 | 0 | 1 | NA | 3 | NA | NA |

| NSD | PPMI (cohort at enrollment) | | | | | | | | | | | PASADENA | SPARK |
|---|---|---|---|---|---|---|---|---|---|---|---|---|---|
| | **Sporadic PD** (N = 252) | **LRRK2 PD** (N = 20) | **GBA PD** (N = 30) | **SNCA PD** (N = 0) | **PRKN PD** (N = 4) | **RBD** (N = 133) | **Hyposmia** (N = 256) | **LRRK2 NMC** (N = 12) | **GBA NMC** (N = 9) | **SNCA NMC** (N = 0) | **HC** (N = 38) | **Early PD** (N = 249) | **Early PD** (N = 236) |
| **Age/Sex-Expected Lowest Putamen SBR <75%,** n (%) | 240 (98%) | 14 (100%) | 17 (94%) | NA | 4 (100%) | 52 (40%) | 122 (48%) | 2 (18%) | 0 | NA | 1 (3%) | NA | 215 (91%) |
| Missing | 8 | 6 | 12 | NA | 0 | 2 | 1 | 1 | 0 | NA | 5 | NA | 0 |
| **Hoehn and Yahr Stage,** n (%) | | | | | | | | | | | | | |
| 0: Asymptomatic | 0 | 0 | 0 | NA | 0 | NA | NA | NA | NA | NA | NA | NA | 0 |
| 1: Unilateral Involvement only | 74 (29%) | 2 (15%) | 6 (32%) | NA | 1 (25%) | NA | NA | NA | NA | NA | NA | NA | 56 (24%) |
| 2: Bilateral Involvement without impairment of balance | 173 (69%) | 10 (77%) | 11 (58%) | NA | 3 (75%) | NA | NA | NA | NA | NA | NA | NA | 164 (69%) |
| 3: Mild to Moderate Involvement | 4 (2%) | 1 (8%) | 2 (11%) | NA | 0 | NA | NA | NA | NA | NA | NA | NA | 2 (1%) |
| Missing | 1 | 7 | 11 | NA | 0 | NA | NA | NA | NA | NA | NA | NA | 14 (6%) |

* MDS-UPDRS Part III > 4 excluding postural and action tremor.

HC= Healthy controls. MDS-UPDRS=Movement Disorder Society Unified Parkinson's Disease Rating Scale. MoCA=Montreal Cognitive Assessment. NMC=non-manifesting carriers. PD=Parkinson's disease. RBD=REM sleep behavior disorder. SBR= Striatal Binding Ratio. UPSIT= University of Pennsylvania Smell Identification Test

### Supplementary Table 3b.

| NSD | PPMI (cohort at enrollment) | | | | | | | | | | | PASADENA | SPARK |
|---|---|---|---|---|---|---|---|---|---|---|---|---|---|
| | **Sporadic PD** (N = 5) | **LRRK2 PD** (N = 10) | **GBA PD** (N = 9) | **SNCA PD** (N = 20) | **PRKN PD** (N = 0) | **RBD** (N = 0) | **Hyposmia** (N = 1) | **LRRK2 NMC** (N = 4) | **GBA NMC** (N = 0) | **SNCA NMC** (N = 7) | **HC** (N = 0) | **Early PD** (N = 0) | **Early PD** (N = 0) |
| **Age (Years),** Mean (SD) | 70.1 (9.0) | 63.2 (6.0) | 64.5 (8.7) | 51.6 (10.9) | NA | NA | 68.5 (NA) | 61.5 (10.9) | NA | 43.9 (15.4) | NA | NA | NA |
| **Sex,** n (%) | | | | | | | | | | | | | |
| Male | 2 (40%) | 5 (50%) | 4 (44%) | 9 (45%) | NA | NA | 1 (100%) | 1 (25%) | NA | 1 (14%) | NA | NA | NA |
| Female | 3 (60%) | 5 (50%) | 5 (56%) | 11 (55%) | NA | NA | 0 | 3 (75%) | NA | 6 (86%) | NA | NA | NA |
| **Years Since PD Diagnosis,** Mean (SD) | 0.8 (0.6) | 3.3 (1.8) | 4.2 (2.1) | 3.5 (3.2) | NA | NA | NA | NA | NA | NA | NA | NA | NA |
| **MDS-UPDRS Item 1.1 Score,** n (%) | | | | | | | | | | | | | |
| 0: Normal | 4 (80%) | 6 (75%) | 8 (89%) | 14 (70%) | NA | NA | 1 (100%) | 3 (75%) | NA | 6 (86%) | NA | NA | NA |
| 1: Slight | 1 (20%) | 2 (25%) | 1 (11%) | 4 (20%) | NA | NA | 0 | 1 (25%) | NA | 1 (14%) | NA | NA | NA |
| 2 - 4: Mild to Severe | 0 | 0 | 0 | 2 (10%) | NA | NA | 0 | 0 | NA | 0 | NA | NA | NA |
| Missing | 0 | 2 | 0 | 0 | NA | NA | 0 | 0 | NA | 0 | NA | NA | NA |
| **MDS-UPDRS Part I,** Mean (SD) | 14.5 (0.7) | 6.0 (3.6) | 4.8 (3.6) | 9.3 (6.6) | NA | NA | - | 4.0 (2.7) | NA | 3.0 (2.3) | NA | NA | NA |
| Missing | 3 | 2 | 0 | 0 | NA | NA | 1 | 0 | NA | 0 | NA | NA | NA |
| **MDS-UPDRS Part II,** Mean (SD) | 5.0 (2.0) | 6.7 (2.8) | 7.9 (6.1) | 8.9 (6.8) | NA | NA | - | 1.0 (1.4) | NA | 0.9 (1.5) | NA | NA | NA |
| Missing | 2 | 1 | 0 | 0 | NA | NA | 1 | 0 | NA | 0 | NA | NA | NA |
| **MDS-UPDRS Part III (OFF),** Mean (SD) | 21.2 (5.3) | 23.3 (11.8) | 22.8 (14.3) | 26.9 (17.9) | NA | NA | 10.0 (NA) | 5.3 (7.2) | NA | 0.2 (0.4) | NA | NA | NA |
| Missing | 0 | 7 | 5 | 2 | NA | NA | 0 | 0 | NA | 1 | NA | NA | NA |
| **Subthreshold Parkinsonism*,** n (%) | 5 (100%) | 3 (100%) | 4 (100%) | 18 (100%) | NA | NA | 1 (100%) | 1 (25%) | NA | 0 | NA | NA | NA |
| Missing | 0 | 7 | 5 | 2 | NA | NA | 0 | 0 | NA | 1 | NA | NA | NA |
| **MOCA Total Score,** Mean (SD) | 27.2 (2.5) | 26.3 (2.7) | 24.8 (4.7) | 25.1 (5.1) | NA | NA | 30.0 (NA) | 28.0 (2.8) | NA | 26.4 (3.2) | NA | NA | NA |
| **On PD Treatment,** n (%) | 0 | 9 (90%) | 7 (78%) | 18 (90%) | NA | NA | 0 | 0 | NA | 0 | NA | NA | NA |
| Missing | 0 | 0 | 0 | 0 | NA | NA | 0 | 0 | NA | 1 | NA | NA | NA |
| **UPSIT Percentile ≤ 15,** n (%) | 5 (100%) | 9 (90%) | 9 (100%) | 19 (100%) | NA | NA | 1 (100%) | 3 (75%) | NA | 4 (67%) | NA | NA | NA |
| Missing | 0 | 0 | 0 | 1 | NA | NA | 0 | 0 | NA | 1 | NA | NA | NA |
| **Age/Sex-Expected Lowest Putamen SBR <75%,** n (%) | 4 (100%) | 2 (100%) | 1 (100%) | 7 (100%) | NA | NA | 1 (100%) | - | NA | 0 | NA | NA | NA |
| Missing | 1 | 8 | 8 | 13 | NA | NA | 0 | 4 | NA | 2 | NA | NA | NA |

| NSD | PPMI (cohort at enrollment) Sporadic PD (N = 5) | LRRK2 PD (N = 10) | GBA PD (N = 9) | SNCA PD (N = 20) | PRKN PD (N = 0) | RBD (N = 0) | Hyposmia (N = 1) | LRRK2 NMC (N = 4) | GBA NMC (N = 0) | SNCA NMC (N = 7) | HC (N = 0) | PASADENA Early PD (N = 0) | SPARK Early PD (N = 0) |
|---|---|---|---|---|---|---|---|---|---|---|---|---|---|
| **Hoeh and Yahr Stage,** n (%) | | | | | | | | | | | | | |
| 0: Asymptomatic | 0 | 0 | 0 | 0 | NA | NA | NA | NA | NA | NA | NA | NA | NA |
| 1: Unilateral Involvement only | 3 (60%) | 1 (33%) | 1 (25%) | 7 (39%) | NA | NA | NA | NA | NA | NA | NA | NA | NA |
| 2: Bilateral Involvement without impairment of balance | 2 (40%) | 2 (67%) | 1 (25%) | 7 (39%) | NA | NA | NA | NA | NA | NA | NA | NA | NA |
| 3: Mild to Moderate Involvement | 0 | 0 | 2 (50%) | 4 (22%) | NA | NA | NA | NA | NA | NA | NA | NA | NA |
| Missing | 0 | 7 | 5 | 2 | NA | NA | NA | NA | NA | NA | NA | NA | NA |

* MDS-UPDRS Part III > 4 excluding postural and action tremor.
HC= Healthy controls. MDS-UPDRS=Movement Disorder Society Unified Parkinson's Disease Rating Scale. MoCA=Montreal Cognitive Assessment. PD=Parkinson's disease. NMC=non-manifesting carriers. RBD=REM sleep behavior disorder. SBR= Striatal Binding Ratio. UPSIT= University of Pennsylvania Smell Identification Test

Supplementary Figure 1. Distribution of NSD stage at study enrollment among early PD participants in the PPMI, PASADENA, and SPARK cohorts with a positive cerebrospinal fluid alpha-synuclein seed amplification assay test result.

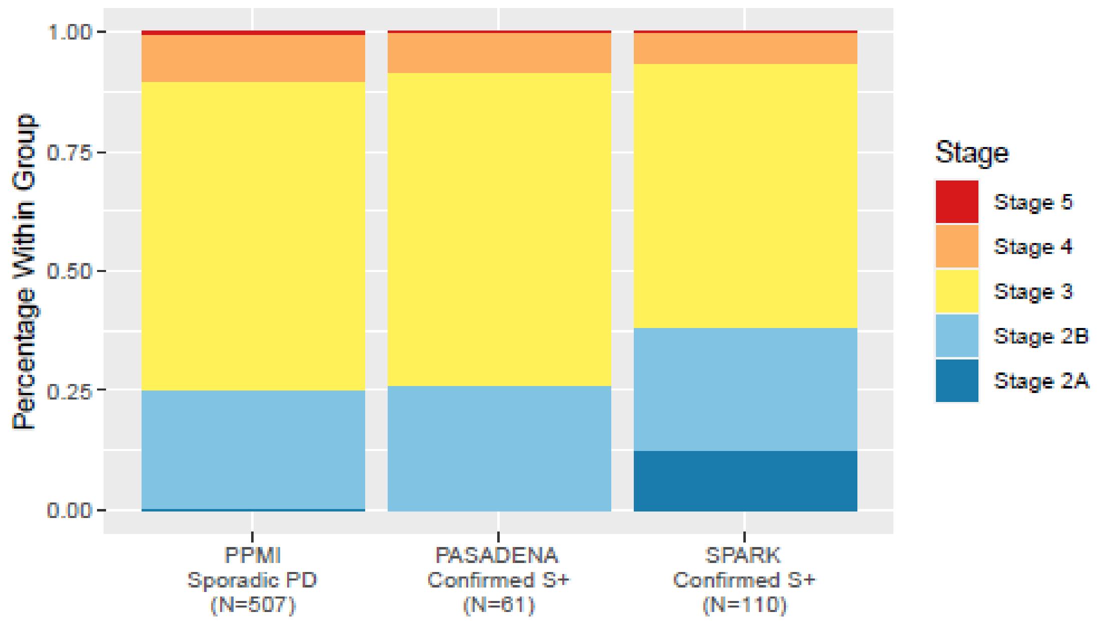


| Stage | PPMI Sporadic PD (N=507) | PASADENA Confirmed S+ (N=61) | SPARK Confirmed S+ (N=110) |
|---|---|---|---|
| Stage 2A | 2 (<1%) | 0 | 14 (13%) |
| Stage 2B | 126 (25%) | 16 (26%) | 28 (25%) |
| Stage 3 | 328 (65%) | 40 (66%) | 61 (55%) |
| Stage 4 | 50 (10%) | 5 (8%) | 7 (6%) |
| Stage 5 | 1 (<1%) | 0 | 0 |
